# Comparative Analysis of COVID-19 Serological testing methods to Polymerase Chain Reaction: A systematic review and meta-analysis

**DOI:** 10.1101/2024.04.18.24305918

**Authors:** David Chisompola, Alex Maleti, Kingsley Tembo, Tioni Banda, George Chishinji, Richard Phiri

## Abstract

**Background:** Accurate and reliable diagnosis of COVID-19 is essential for effective disease management and public health interventions. SARS-CoV-2 antibody/antigen tests play a crucial role in identifying covid-19 infections and assessing immunity in populations. This systematic review aimed to evaluate the sensitivity, specificity, and accuracy of SARS-CoV-2 antibody tests in diagnosing COVID-19.

**Method:** A comprehensive systematic review of the literature was conducted using prominent scientific databases, including PubMed, Google Scholar, Cochrane, and Scopus to search studies published from January 2020 to May 2023. The review followed the PRISMA-DTA framework to ensure transparency and rigor in the selection and evaluation of studies. The QUADAS-2 tool was utilized to assess study quality and risk of bias. This review was registered on PROSPERO (registration number CRD 42023445695).

**Results:** The pooled sensitivity of SARS-CoV-2 antibody/antigen tests was found to be 73% (95% CI: 60–86), with individual studies reporting a wide range from 30% to 100%. The pooled specificity was 98% (95% CI: 97–100), with values ranging from 85.9% to 100%. The pooled accuracy was 88%, showing variation from 50% to 100% across different studies.

**Conclusion:** This review highlights the moderate to high sensitivity and specificity of SARS-CoV-2 antibody/antigen tests for COVID-19 diagnosis. The substantial variability in test performance necessitates the standardization of testing protocols and further research to improve accuracy and reliability. These findings offer valuable insights for clinical decision-making and the formulation of effective public health strategies related to COVID-19 diagnosis.

## Introduction

The global outbreak of the novel coronavirus disease (COVID-19), caused by severe acute respiratory syndrome coronavirus 2 (SARS-CoV-2), has led to an unprecedented public health crisis, affecting millions of lives and economies worldwide ^1^. According to world health organization (WHO) estimates, the COVID-19 pandemic has affected over 200 countries and territories, with 614,385,693 confirmed cases and an additionally 6,522,600 COVID-19 associated deaths as of 2022 ^2^.

Since the emergence of SARS-CoV-2, efforts to understand, control, and mitigate its spread have been of paramount importance. Central to these efforts, has been the availability, accuracy and timely detection of SARS-CoV-2 infections, which has driven the development of various diagnostic techniques ^3^. Among the diagnostic methods utilized for COVID-19 detection, two primary approaches have emerged: polymerase chain reaction (PCR) and serological testing ^4^. PCR is a molecular technique used to directly detect viral genetic material in respiratory samples, enabling the diagnosis of active infections. On the other hand, serological tests detect specific antibodies produced by the immune system in response to the virus symptomatic or asymptomatic individuals, providing insights into the individual’s past exposure and immune response ^4^.

As the pandemic has evolved, an increasing array of serological testing methods and PCR-based assays have been developed and deployed for clinical and epidemiological purposes. These methods vary in terms of sensitivity, specificity, ease of use, and turnaround time, each with its own strengths and limitations. Given the diversity of available testing strategies, there is a growing need to comprehensively compare and evaluate their performance characteristics ^4^. Such a comparative analysis is crucial for informing healthcare decision-makers, guiding testing strategies, and understanding the limitations and advantages of different diagnostic approaches ^5^.

Diagnostic accuracy assessment of Polymerase Chain Reaction (PCR) and serological testing for SARS-CoV-2 infection has been the focus of numerous studies, examining key parameters like sensitivity, specificity, positive predictive value (PPV), and negative predictive value (NPV). While individual studies have explored the precision and utility of COVID-19 serological tests and PCR assays, a comprehensive evaluation of diagnostic accuracy for both methods have been achieved through quantitative data pooling across studies. This approach yields more robust estimates of test performance, providing a comprehensive synthesis of existing evidence.

Considering PCR testing, some studies have reported high sensitivity and specificity, while others highlighted challenges such as false-negative results due to sampling errors or viral load variations. Serological testing also exhibited variability, influenced by factors like sample collection timing, antibody kinetics, and cross-reactivity. Collectively, these studies yield insights into serological tests and Ag-RDTs’ diagnostic performance for detecting SARS-CoV-2 infection. While sensitivity varies, specificity remains consistently high, underscoring the need to consider performance limitations when using these tests for COVID-19 diagnosis.

Understanding comparative accuracy between PCR and serological testing is vital for optimizing diagnostic strategies, especially in resource-limited settings. Factors like turnaround time, cost, and platform availability impact diagnostic method selection. Therefore, this research aims to systematically review and meta-analyze COVID-19 serological testing methods and PCR assays. By synthesizing diverse study data, the study aims to comprehensively outline each approach’s strengths and limitations, facilitate evidence-based decision-making, and enhance understanding of SARS-CoV-2 infection dynamics.

This systematic review intends to comprehensively evaluate Serological testing accuracy for COVID-19 diagnosis relative to PCR testing. Synthesizing evidence and appraising study quality, it will compare pooled sensitivities and specificities of different methods, aiding evidence-based clinical practice and public health strategies. The comparative analysis of COVID-19 serological testing to PCR via systematic review and meta-analysis promises to enhance diagnostic understanding. The study’s outcomes are expected to impact clinical practice, epidemiology, and public health responses to the ongoing pandemic.

## Methods

The systematic review protocol was registered on PROSPERO (CRD42023445695) and was conducted in accordance with the Preferred Reporting Items for Systematic Reviews and Meta-Analyses (PRISMA) reporting guidelines ^8^.

### Search strategy and selection

A literature search was conducted in electronic databases, including PubMed, Google Scholar, Scopus, and Cochrane, to identify relevant studies. The search strategy was developed using appropriate Medical Subject Headings (MeSH) terms and keywords related to COVID-19, PCR testing, serology testing, diagnostic accuracy, sensitivity and specificity ^6^. Boolean operators (AND, OR) were used to combine search terms effectively. The search strategy was tailored to the requirements of each database ^6^. Studies involving individuals suspected or confirmed to have COVID-19 was included. No age, gender, or geographical restrictions was applied ^6^. Two independent reviewers screened the titles and abstracts of retrieved articles and identified potentially relevant studies published from January 2020 to May 2023. Full texts of the selected articles were obtained and assessed against the inclusion and exclusion criteria. Any discrepancies between the reviewers were resolved through discussion or consultation with a third reviewer. The following two strategies were used to search for articles:

#1: “Severe Acute Respiratory Syndrome Coronavirus 2”[Supplementary Concept] OR “COVID-19”[Supplementary Concept] OR “Betacoronavirus”[MeSH Terms] OR “Coronavirus”[MeSH Terms] OR “covid*”[Text Word] OR “coronavirus*”[Text Word] OR “corona virus*”[Text Word] OR “ncov*”[Text Word] OR “n cov*”[Text Word] OR “sarscov*”[Text Word] OR “sars cov*”[Text Word] OR “2019ncov*”[Text Word] OR “2019 ncov*”[Text Word] OR “sars2*”[Text Word] OR “sars 2*”[Text Word]

#2: “covid-19 testing”[MeSH Terms] OR “Antigen/Antibody based rapid diagnostic test”[All Fields] OR “diagnostic test” OR “Rapid antigen” OR “lateral flow antigen detection” OR “lateral flow assay*” OR “ Serological Testing”[Mesh].

### Eligibility criteria

The criteria for including or excluding studies, which were used to decide if screened studies were suitable for review, are outlined below.

### Inclusion

- Original research studies, including randomized controlled trials, cohort studies, case-control studies, cross-sectional studies, and observational studies, that reported the sensitivity, specificity, and accuracy of SARS-CoV-2 antibody tests.
- Both men and women of any age were considered.
- Subjects had to have received a COVID-19 diagnosis.
- Studies in languages other than English were accepted if they had an English translation available in full text.

### Exclusion

- Excluded were qualitative analyses, reviews of existing literature, opinion pieces, and official documents related to policies.
- Research focused on patients with infections other than COVID-19 was not considered.
- Studies lacking the desired outcome (i.e., Rapid COVID-19 diagnosis where PCR was not used as an index test) were omitted.
- Studies lacking complete textual content were not included.

### Data extraction and synthesis

Data was independently extracted from selected studies by using a developed Microsoft excel sheet. The following data was extracted: general study details (authors, year of publication, country and sample size), methods, characteristics, and diagnostic test results (Sensitivity, Specificity, and Accuracy) ^6^ from 1 January 2020 to 23 May 2023. Two researchers were involved in the process of quality assessment. To evaluate the risk of bias and applicability of the included studies the “Quality Assessment of Diagnostic Accuracy Studies 2” (QUADAS-2) ^7^ tool was used.

### Quality assessment

We performed a meta-analysis on pooled studies, Sensitivity, specificity and accuracy of calculated estimates for each included study. Statistical analysis was performed using Statistical Package for the Social Sciences (SPSS) and Review Manager (RevMan) 5.4.1. We used already calculated estimate at 95% confidence interval of pooled sensitivity and pooled specificity data ^6^. The included studies were analysed on variations or inconsistencies in findings using random-effects meta-analysis. Heterogeneity was assessed using summary receiver operating characteristic (ROC) curves with 95% prediction regions estimated using bivariate meta-analysis with test-level random effect, and forest plots ^8^.

## Results

A total of 353 articles were initially screened for inclusion, and ultimately, 13 articles were selected for analysis. These 13 articles, encompassing a total sample size of 3759 participants.

**Figure 1.**
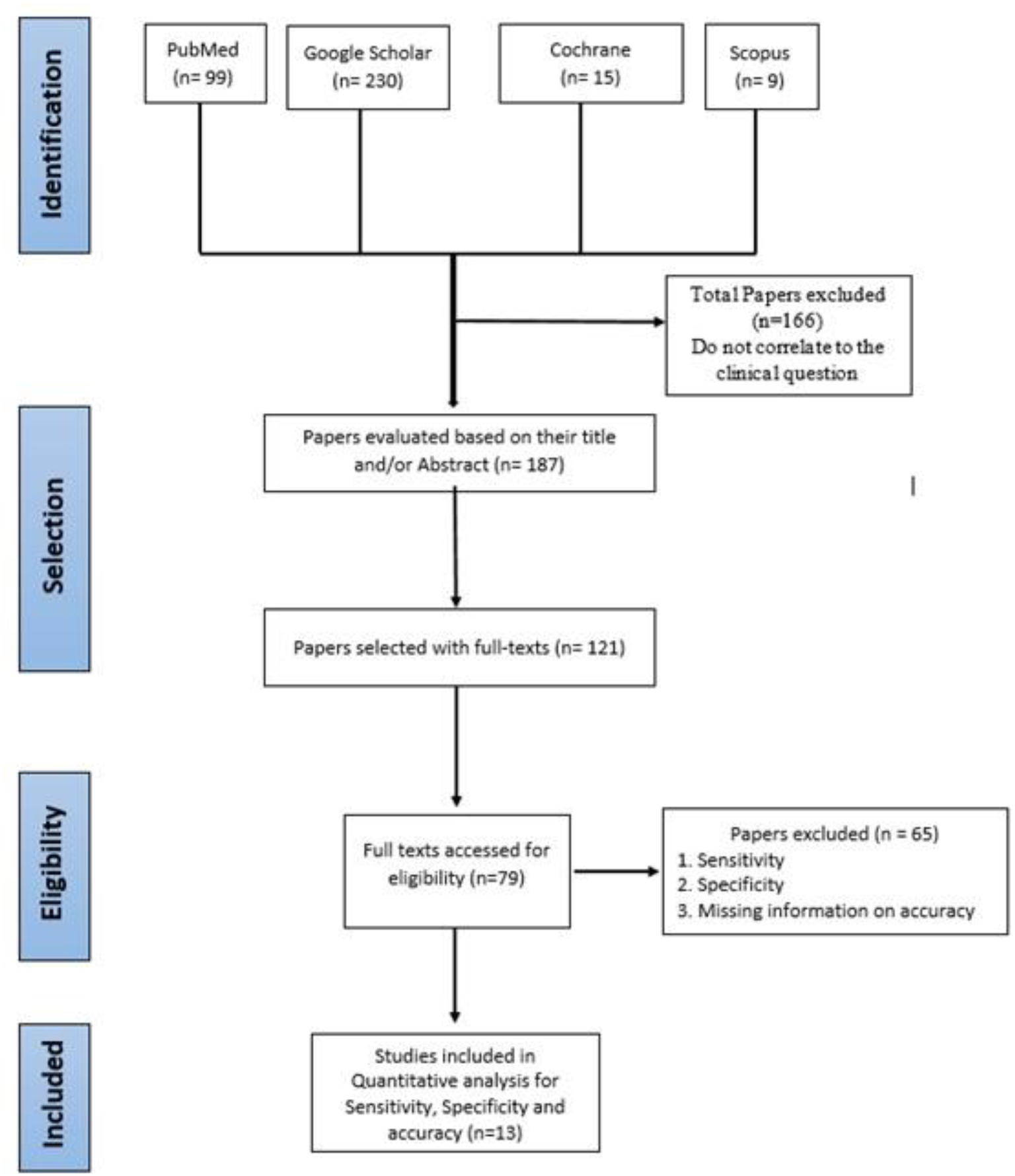
presents the PRISMA flow diagram of the conducted study.

**Figure 2.**
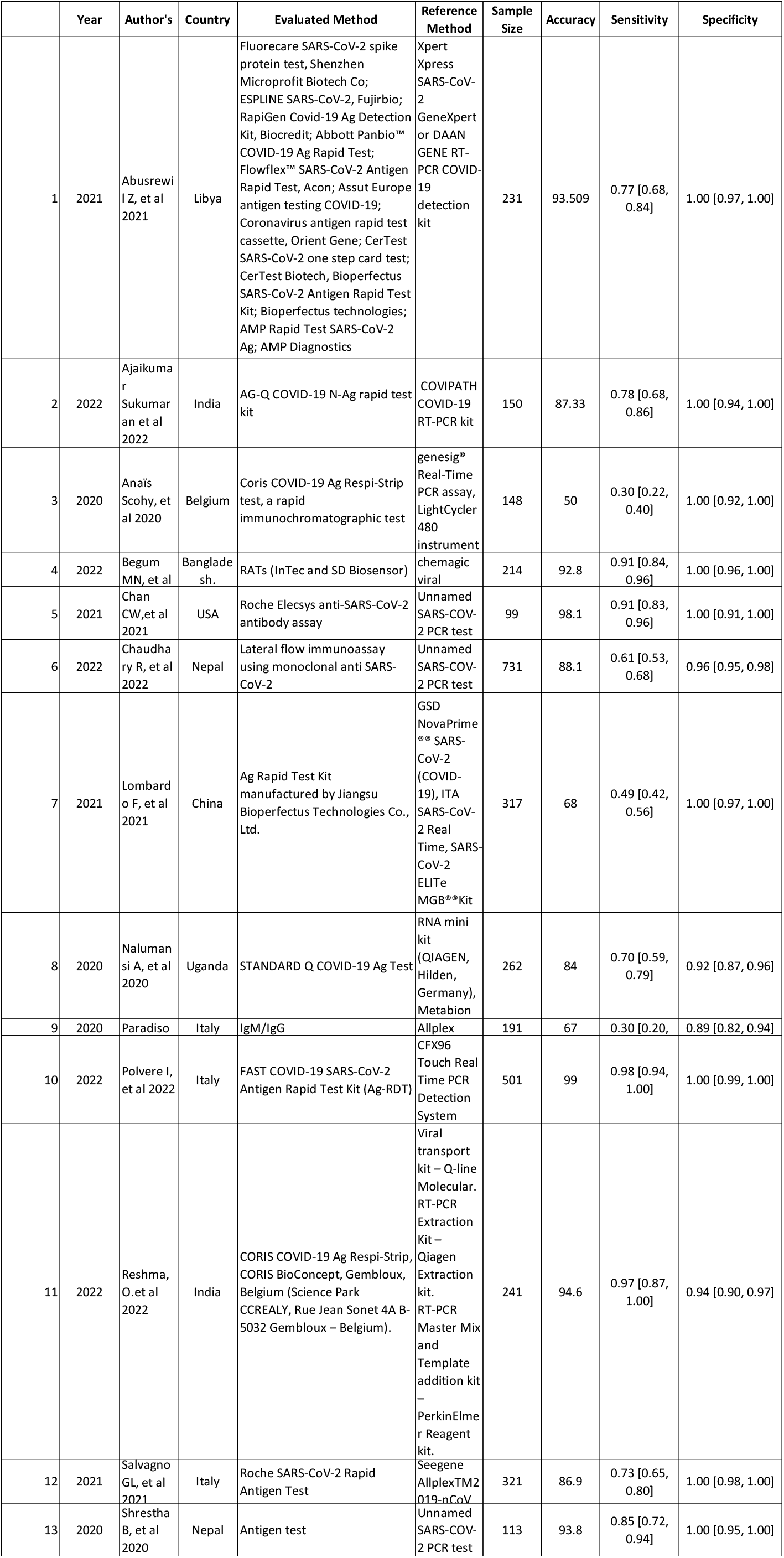
shows Characteristics of the included studies.

QUADAS-2 risk of bias and applicability concerns summary review authors’ judgment about each domain for 13 test evaluations reported in 13 included studies.

**Figure 3 and 4.**
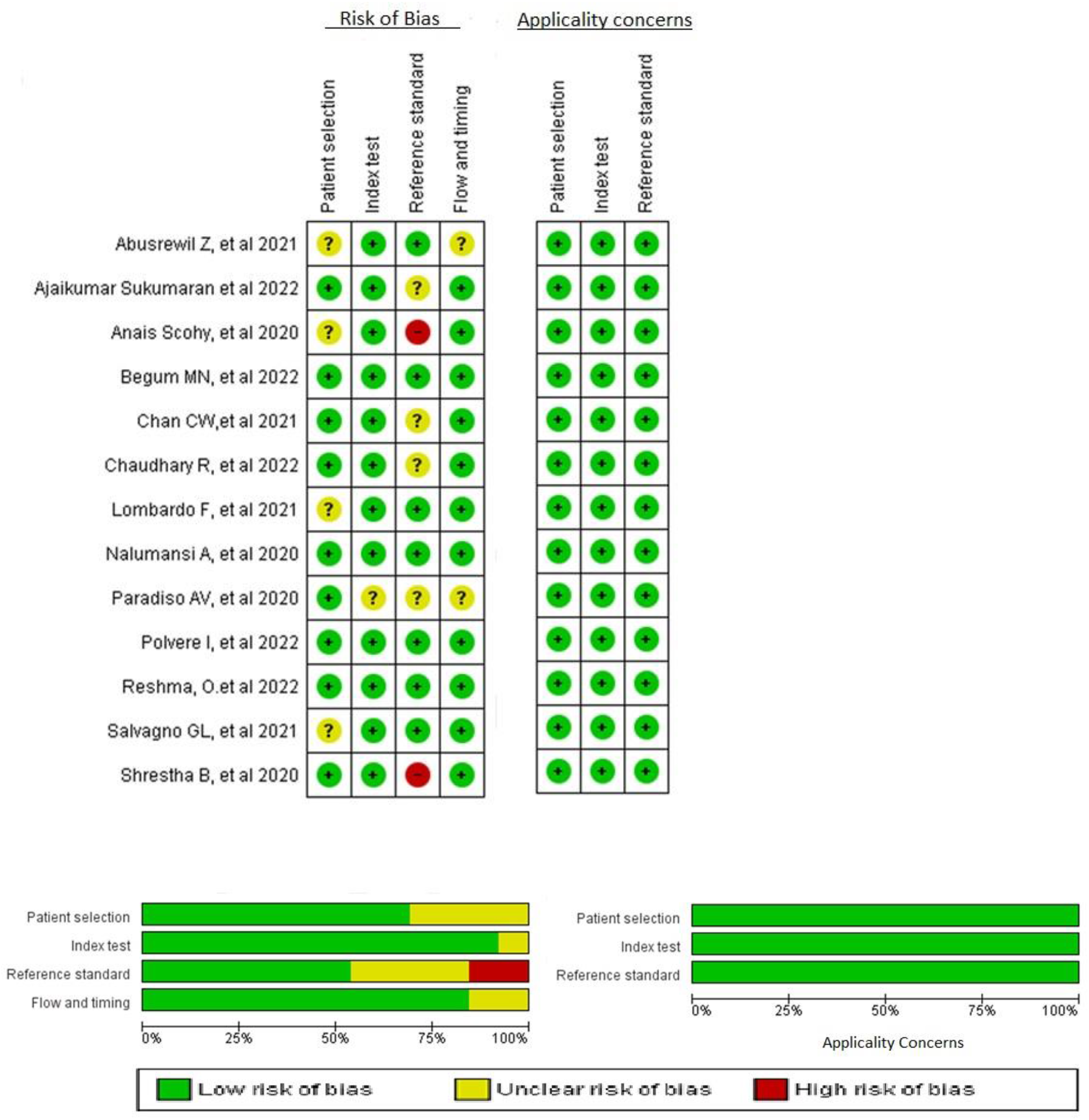
Shows QUADAS-2 quality assessment results.

In this analysis, the term ‘Unclear’ is not indicative of a ‘medium’ risk of concern; rather, it signifies a lack of adequate information, making it impossible to categorize the study as either at ‘low’ risk of bias or inapplicable ^9^.

**Figure 5.**
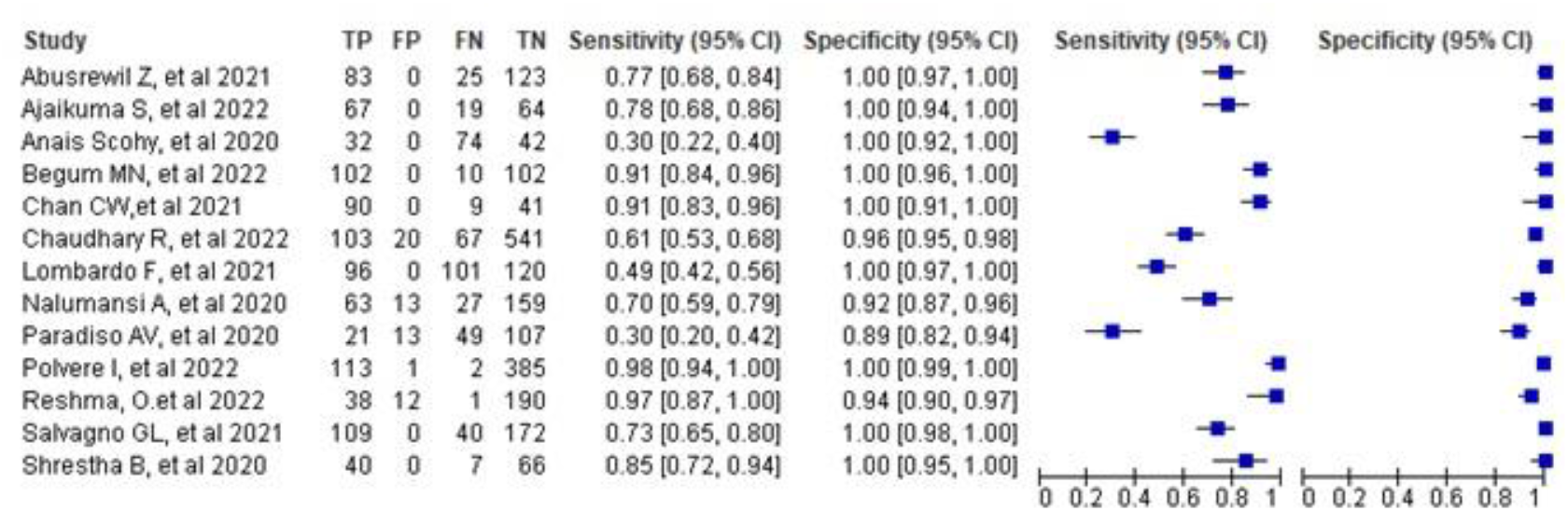
shows overall Meta-Analysis for Sensitivity and Specificity of RDTs.

The overall pooled sensitivity and specificity of 13 studies was 73 (95% CI: 60–86) and 98 (95% CI: 97–100)

**Figure 6.**
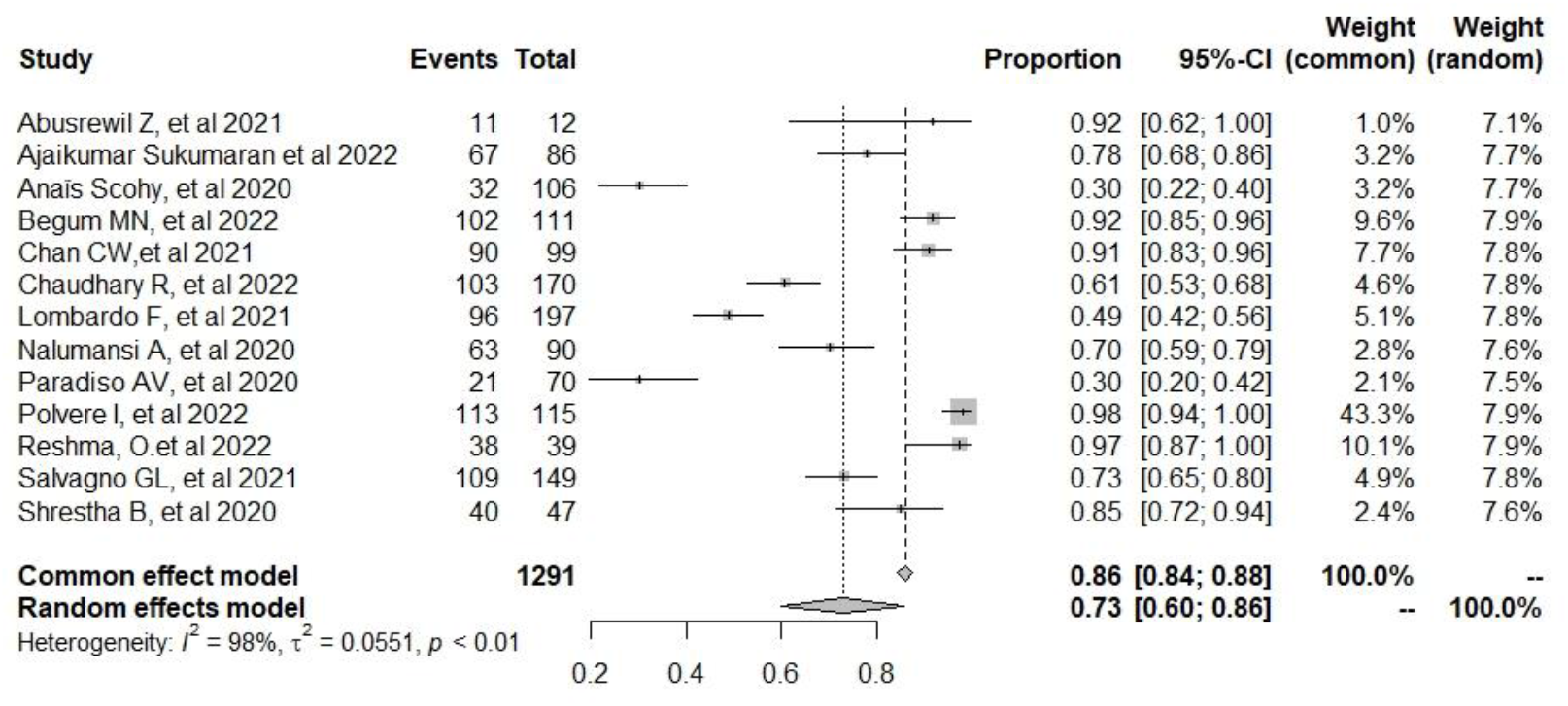
shows Meta-Analysis for Sensitivity of RDTs.

**Figure 7.**
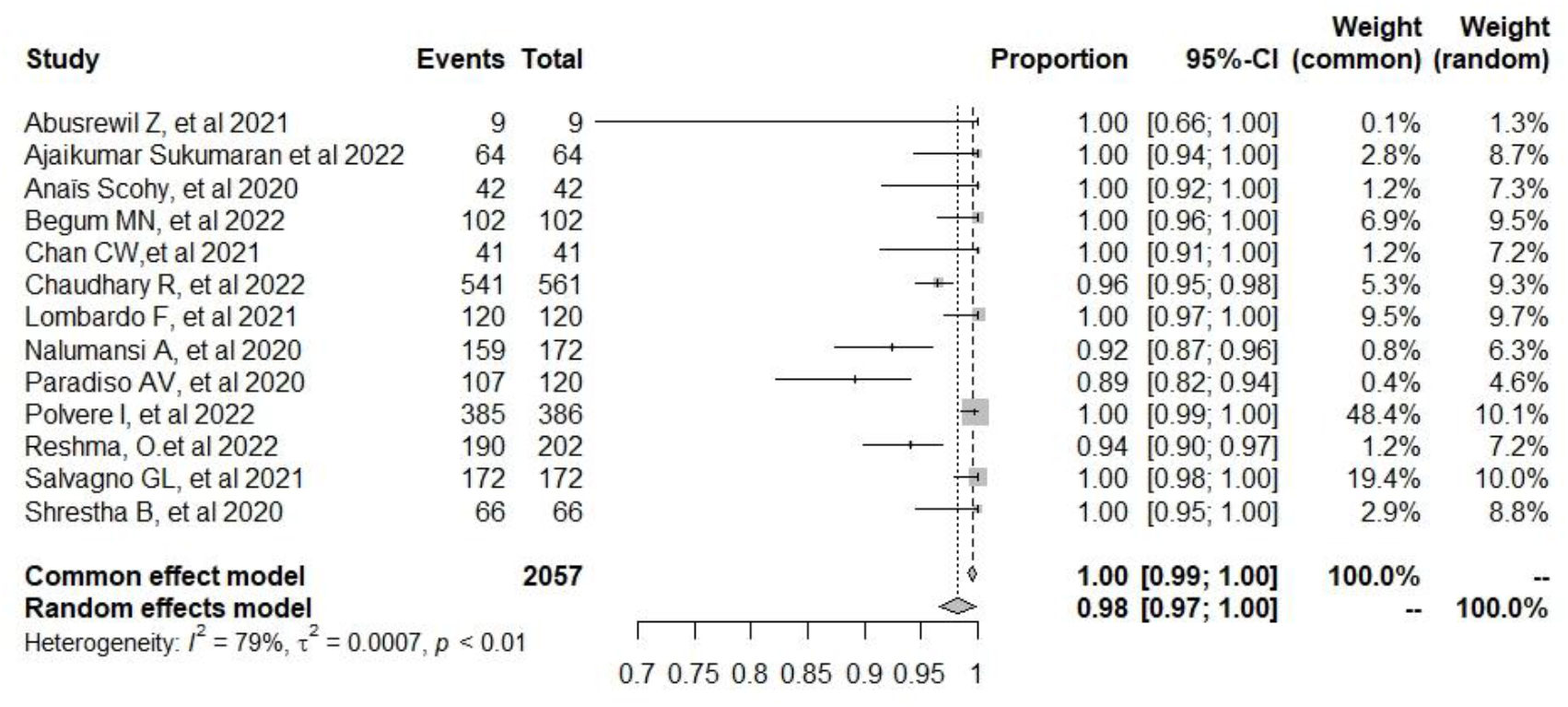
shows Meta-Analysis for Specificity of RDTs.

**Figure 8.**
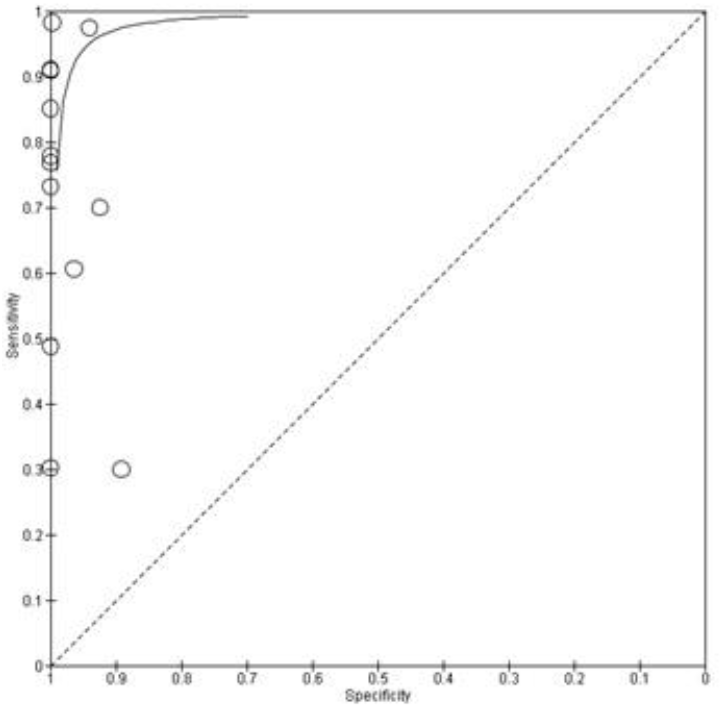
Shows the Receiver Operating Characteristic curve.

## Discussion

The accurate and reliable detection of SARS-CoV-2 infection is critical for effective disease management and public health interventions. In this study, we focused on evaluating the diagnostic accuracy, sensitivity, specificity, and overall performance of SARS-CoV-2 rapid tests. Literature provides valuable insights into the effectiveness of RDT’s in various populations. 13 studies from diverse geographical regions, including Bangladesh, Belgium, China, Italy, India, Libya, Nepal, and the USA were included, highlighting its global relevance. The inclusivity of studies from Asia, Africa, Europe, and North America further emphasizes the broad scope of the study’s analysis.

Meta-analysis, incorporated data from 13 studies ^10,11,20,21,12–19^, specifically examined the diagnostic accuracy of RDTs. The pooled sensitivity of RDTs across all studies was observed to be 73% (95% CI: 60–86), which is notably below the minimum performance requirement established by WHO at ≥ 80% ^22^. This finding underscores a significant challenge in the overall sensitivity of these tests, potentially impacting their effectiveness in accurately identifying individuals with SARS-CoV-2 infection. Sensitivity is a crucial metric as it reflects the ability of a test to correctly identify true positive cases and falling below the WHO threshold may indicate a risk of false-negative results.

Conversely, the specificity of the RDTs proved to be robust, surpassing the WHO cut-off at an impressive 98% (95% CI: 97–100). Specificity gauges the ability of a test to correctly identify true negative cases, and the high specificity observed in this analysis indicates a low rate of false positives. A specificity of 98% suggests that these tests are generally accurate in correctly identifying individuals without SARS-CoV-2 infection, reducing the likelihood of unnecessary anxiety or further diagnostic procedures for individuals who do not have the virus.

It is important to note the variability in the performance of the RDTs across different studies. For instance, a study conducted by Juan Jeferson Vilca-Alosilla et *al*. in 2023 reported a slightly higher median sensitivity of 94.5% and a specificity of 98.4%. This variability suggests that factors such as RDTs brand used, study design, population characteristics, and regional differences may influence the performance outcomes of these tests. Moreover, it is also important to emphasize the need for continuous monitoring and evaluation of diagnostic tools to ensure their reliability across diverse settings. In contrast Pandey et *al*.’s study in 2022 provides additional insights, reporting a pooled sensitivity of 78.2% and an impressive specificity of 99.5%. While the sensitivity remains below the WHO threshold, the high specificity suggests that RDTs are effective in correctly identifying individuals without SARS-CoV-2 infection. These nuanced variations in sensitivity and specificity highlight the complexity of evaluating diagnostic tools and the importance of considering multiple studies to derive a comprehensive understanding of their performance characteristics.

Vilca-Alosilla JJ et *al* 2023, examined and unveiled a wide spectrum of diagnostic sensitivity, ranging from 36.8% to 99.2%, with a median sensitivity of 94.5%. Similarly, the specificity of serological tests displayed variability, spanning from 79.3% to 99.8%, and a median specificity of 98.4% ^22^. These findings underscore the significant diversity in sensitivity and specificity values observed across various studies, emphasizing the variability in the performance of serological tests for SARS-CoV-2 detection.

Additionally, Fujita-Rohwerder N et *al* 2022, states that there was an elevated pooled estimates of sensitivity and specificity for serological testing with a pooled sensitivity of 64.2% (95% CI 57.4% to 70.5%) and a pooled specificity of 99.1% (95% CI 98.2% to 99.5%) ^23^. This outcome suggests a moderate level of sensitivity while indicating a notably high specificity when utilizing serological tests to detect SARS-CoV-2 infection. Furthermore, in a study done by Brummer LE et *al* 2023, explored rapid diagnostic tests for SARS-CoV-2 detection, finding a moderate sensitivity (71.2%, 95% CI 68.2% to 74.0%) and high specificity (98.9%, 95% CI 98.6% to 99.1%)^7^.

It is crucial to note that while Reverse Transcription-Quantitative PCR (RT-qPCR) is widely accepted for COVID-19 diagnosis, its limitations, such as the cost, requirement for specialized equipment and centralized testing facilities, hinder its accessibility in local clinics. To address this, RDTs have been approved as an alternative, as they do not rely on expensive machinery and can provide quicker results. Additionally, the need to transport samples to locations with RT-qPCR capabilities introduces delays and adds to the anxiety of individuals awaiting test results. In as much as RDT kits are cheaper than PCR techniques, it is critical that true positive and true negatives are correctly detected by these testing kits. A low sensitivity performance analysed by this present study is suggestive of possibly missing positive covid-19 cases thereby misdiagnosing individuals with covid-19.

Despite the challenges posed by varying sensitivity levels, the findings underscore the significance of continuous evaluation and improvement in the performance of SARS-CoV-2 diagnostic tests. As the global landscape of the COVID-19 pandemic evolves, the findings contribute to the ongoing efforts to optimize testing strategies, enhance accuracy, and ultimately facilitate more effective disease management and public health interventions.

Emanating from the evaluation of study quality using QUADAS-2, most of the included research displayed a low level of bias. However, a few studies highlighted a heightened risk of bias in QUADAS-2 assessment. The uncertainty in patient selection risk arose from insufficient information regarding the criteria for including or excluding patients. Some studies lacked clarity regarding the risk associated with the index test, as authors did not specify whether they interpreted index test results while aware of reference test outcomes. Additionally, the high risk of bias linked to the reference standard was due to the interpretation of results without considering the index test outcomes.

A total of 3759 research participants from the 13 included studies examined the efficacy of antigen tests using 24 different test kits against the RT-PCR reference standard. The majority of research was published in 2022 (n=5), 2021 (n=4), and 2020 (n=4). Numerous studies that used articles from nations including Italy (n = 3), India (n = 2), Nepal (n = 2), USA (n = 1), China (n = 1), Belgium (n = 1), Bangladesh (n = 1), Uganda (n=1), and Libya (n=1) reported on the effectiveness of RDTs for SARS-CoV-2. Test kits included the following: CerTest SARS-CoV-2 one step card test, Bioperfectus SARS-CoV-2 Antigen Rapid Test Kit, Roche SARS-CoV-2 Rapid Antigen Test, Roche Elecsys anti-SARS-CoV-2 antibody assay, RapiGen Covid-19 Ag Detection Kit, Abbott PanbioTM COVID-19 Ag Rapid Test, Assut Europe antigen testing COVID-19, RATs (InTec and SD Biosensor), CORIS COVID-19 Ag Respi-Strip, Coris BioConcept COVID 19 Ag Respi-Strip, Lateral flow immunoassay, Maglumi (Snibe), Liaison (Diasorin), iFlash (Yhlo), Euroimmun (Medizinische Labordiagnostika AG), IgM/IgG (VivaDiag), AG-Q COVID-19 N-Ag rapid test kit, Coris COVID-19 Ag Respi-Strip test, a rapid immunochromatographic test, Ag Rapid Test Kit manufactured by Jiangsu Bioperfectus Technologies Co., Ltd., Antigen test (Un-named), FAST COVID-19 SARS-CoV-2 Ag-RDT, Flowflex™ SARS-CoV-2 Antigen Rapid Test, AMP Rapid Test SARS-CoV-2 Ag, and Wantai (Wantai Biological Pharmacy) assays.

The examination of study quality using the QUADAS-2 tool unveiled considerable variability across all four domains. Concerning individual selection, a majority of studies (69.2%, n=9) were designated as having a low risk of bias, denoting a clear and well-defined participant selection process. However, a notable fraction of studies (30.8%, n=4) fell into the ‘unclear’ risk of bias category, indicating insufficient reporting on participant selection methods, such as the enrollment of a consecutive or random sample.

Regarding the reference standard, studies predominantly utilized RT-PCR as the gold standard. A small proportion of studies (15.4%, n=2) were identified as having a high risk of bias due to inadequate reporting of blinding procedures, potentially introducing biases in the comparison with the gold standard.

Conversely, the risk of bias in the flow and timing domain was low in the majority of studies (84.6%, n=11). These studies explicitly outlined the inclusion of both asymptomatic and symptomatic patients seeking SARS-CoV-2 testing. These findings underscore the imperative for improved reporting and adherence to methodological standards in future studies, aiming to enhance the overall quality of research in this domain.

The limitation of this study is that the diagnostic accuracy assessment relies on the available literature, which may exhibit inherent variability due to differences in study designs, sample populations, testing protocols, and reporting practices. Additionally, we had no restrictions on the RDT’s, and the reference method used in this review, the limited number of studies and potential heterogeneity in terms of assay types and outcome measures might restrict the scope for comprehensive meta-analysis, potentially impacting the ability to provide a definitive synthesis of the performance of COVID-19 serological testing methods compared to PCR in detecting SARS-CoV-2 infection.

## Conclusion

In conclusion the comprehensive review of the literature on the diagnostic accuracy, sensitivity, and specificity of SARS-CoV-2 rapid tests provides important insights into their performance and implications for COVID-19 diagnosis. It is important to consider the limitations and heterogeneity observed among the included studies. Variations in test methodologies, sample populations, and the timing of antibody detection contribute to the observed variability in sensitivity, specificity, and accuracy. Standardization of testing protocols and further research efforts are needed to address these limitations and improve the consistency and reliability of SARS-CoV-2 rapid testing. The findings of this review have significant implications for the diagnosis and management of COVID-19 infection. SARS-CoV-2 antibody/antigen tests can play a valuable role in identifying individuals who have been previously infected or have developed immunity to the virus. Refining the accuracy of SARS-CoV-2 antibody/antigen rapid testing kits can enhance the ability to control the spread of the virus, effectively manage COVID-19 cases, and make informed decisions for the well-being of individuals and communities worldwide.

## Data Availability

All data produced or analyzed in the study is provided in this article. Additional related data can be requested from the corresponding author.

## Declaration

All authors have no conflict of interest to declare.

## Ethical approval

The study was conducted under a protocol that was reviewed and approved by PROSPERO using the registration number PROSPERO CRD42023445695.

## Authors’ and contributions

D.C conceptualized the study, conducted the initial analysis and wrote the first draft. D.C conducted the formal analysis and reviewed and edited the final manuscript. T.B, R.P, A.M, K.T, and G.C performed the data curation and reviewed the manuscript. R.P supervised the conduct of the study. All authors have critically reviewed and approved the final draft and are responsible for the content and similarity index of the manuscript

## References

1. Sharma A, Tiwari S, Deb MK, Marty JL. Severe acute respiratory syndrome coronavirus-2 (SARS-CoV-2): a global pandemic and treatment strategies. Int J Antimicrob Agents. 2020 Aug;56(2):106054.

2. Kaul V, Chahal J, Schrarstzhaupt IN, Geduld H, Shen Y, Cecconi MH, et al. Lessons Learned from a Global Perspective of Coronavirus Disease-2019. Clin Chest Med. 2023 Jun;44(2):435–49.

3. Khandia R, Singhal S, Alqahtani T, Kamal MA, El-Shall NA, Nainu F, et al. Emergence of SARS-CoV-2 Omicron (B.1.1.529) variant, salient features, high global health concerns and strategies to counter it amid ongoing COVID-19 pandemic. Environ Res. 2022 Jun;209:112816.

4. Bastos ML, Tavaziva G, Abidi SK, Campbell JR, Haraoui LP, Johnston JC, et al. Diagnostic accuracy of serological tests for covid-19: systematic review and meta-analysis. BMJ [Internet]. 2020 Jul 1;370:m2516. Available from: http://www.bmj.com/content/370/bmj.m2516.abstract

5. La Marca A, Capuzzo M, Paglia T, Roli L, Trenti T, Nelson SM. Testing for SARS-CoV-2 (COVID-19): a systematic review and clinical guide to molecular and serological in-vitro diagnostic assays. Reprod Biomed Online. 2020 Sep;41(3):483–99.

6. Macedo ACL, Prestes G da S, Colonetti T, Candido ACR, Uggioni MLR, Gomes AC, et al. A systematic review and meta-analysis of the accuracy of SARS-COV-2 IGM and IGG tests in individuals with COVID-19. J Clin Virol Off Publ Pan Am Soc Clin Virol. 2022 Mar;148:105121.

7. Brümmer LE, Katzenschlager S, Gaeddert M, Erdmann C, Schmitz S, Bota M, et al. Accuracy of novel antigen rapid diagnostics for SARS-CoV-2: A living systematic review and meta-analysis. PLoS Med. 2021 Aug;18(8):e1003735.

8. Pandey S, Poudel A, Karki D, Thapa J. Diagnostic accuracy of antigen-detection rapid diagnostic tests for diagnosis of COVID-19 in low-and middle-income countries: A systematic review and meta-analysis. PLOS Glob public Heal. 2022;2(4):e0000358.

9. Spick M, Lewis HM, Wilde MJ, Hopley C, Huggett J, Bailey MJ. Systematic review with meta-analysis of diagnostic test accuracy for COVID-19 by mass spectrometry. Metabolism. 2022 Jan;126:154922.

10. Paradiso AV, De Summa S, Loconsole D, Procacci V, Sallustio A, Centrone F, et al. Rapid Serological Assays and SARS-CoV-2 Real-Time Polymerase Chain Reaction Assays for the Detection of SARS-CoV-2: Comparative Study. J Med Internet Res [Internet]. 2020;22(10):e19152. Available from: http://www.jmir.org/2020/10/e19152/

11. Sukumaran A, Suvekbala V R AK, Thomas RE, Raj A, Thomas T, et al. Diagnostic Accuracy of SARS-CoV-2 Nucleocapsid Antigen Self-Test in Comparison to Reverse Transcriptase–Polymerase Chain Reaction. J Appl Lab Med. 2022 Jun 30;7(4):871–80.

12. Scohy A, Anantharajah A, Bodéus M, Kabamba-Mukadi B, Verroken A, Rodriguez-Villalobos H. Low performance of rapid antigen detection test as frontline testing for COVID-19 diagnosis. J Clin Virol. 2020 Aug;129:104455.

13. Lombardo F, Triolo G, Yang B, Liu Z, Maiuri P, Orsini E, et al. Diagnostic Performance of a Rapid Antigen Test Compared with the Reverse Transcription Polymerase Chain Reaction for SARS-CoV-2 Detection in Asymptomatic Individuals Referring to a Drive-in Testing Facility. COVID. 2021 Dec 17;1(4):784–9.

14. Shrestha B, Neupane AK, Pant S, Shrestha A, Bastola A, Rajbhandari B, et al. Sensitivity and Specificity of Lateral Flow Antigen Test Kits for COVID-19 in Asymptomatic Population of Quarantine Centre of Province 3. Kathmandu Univ Med J (KUMJ). 18(70):36–9.

15. Polvere I, Voccola S, D’Andrea S, Zerillo L, Varricchio R, Madera JR, et al. Evaluation of FAST COVID-19 SARS-CoV-2 Antigen Rapid Test Kit for Detection of SARS-CoV-2 in Respiratory Samples from Mildly Symptomatic or Asymptomatic Patients. Diagnostics. 2022 Mar 7;12(3):650.

16. Chaudhary R, Bhatta S, Singh A, Pradhan M, Moktan B, Duwal S, et al. A Comparative Study of Rapid SARS-Cov-2 Antigen Detection Assay against RT-PCR Assay for Diagnosis of COVID-19 in a Tertiary Hospital of Kathmandu. Kathmandu Univ Med J (KUMJ). 2022;20(79):337–41.

17. OR Reshma, Ashna Ajimsha, Ganga Raju Krishna,, Ashish Jitendranath, JT Ramani Bai, KN Deepthi. Evaluation of rapid antigen test against reverse transcription-polymerase chain reaction for the diagnosis of severe acute respiratory syndrome coronavirus 2 in a tertiary care centre in South Kerala.

18. Begum MN, Jubair M, Nahar K, Rahman S, Talha M, Sarker MS, et al. Factors influencing the performance of rapid SARS‐CoV‐2 antigen tests under field condition. J Clin Lab Anal. 2022 Feb 23;36(2).

19. Salvagno GL, Gianfilippi G, Bragantini D, Henry BM, Lippi G. Clinical assessment of the Roche SARS-CoV-2 rapid antigen test. Diagnosis. 2021 Aug 26;8(3):322–6.

20. Chan CW, Shahul S, Coleman C, Tesic V, Parker K, Yeo KTJ. Evaluation of the Truvian Easy Check COVID-19 IgM/IgG Lateral Flow Device for Rapid Anti-SARS-CoV-2 Antibody Detection. Am J Clin Pathol. 2021 Feb 4;155(2):286–95.

21. Abusrewil Z, Alhudiri IM, Kaal HH, El Meshri SE, Ebrahim FO, Dalyoum T, et al. Time scale performance of rapid antigen testing for SARS‐CoV‐2: Evaluation of 10 rapid antigen assays. J Med Virol. 2021 Dec 28;93(12):6512–8.

22. Vilca-Alosilla JJ, Candia-Puma MA, Coronel-Monje K, Goyzueta-Mamani LD, Galdino AS, Machado-de-Ávila RA, et al. A Systematic Review and Meta-Analysis Comparing the Diagnostic Accuracy Tests of COVID-19. Vol. 13, Diagnostics. 2023.

23. Fujita-Rohwerder N, Beckmann L, Zens Y, Verma A. Diagnostic accuracy of rapid point-of-care tests for diagnosis of current SARS-CoV-2 infections in children: a systematic review and meta-analysis. BMJ Evidence-Based Med [Internet]. 2022;27(5):274–87. Available from: https://ebm.bmj.com/content/27/5/274

